# Pharmacist Impact on First-Line Antihypertensives in African American Patients

**DOI:** 10.1101/2024.09.11.24313518

**Authors:** Mia Y. Reid, Jamie E. Coates, Naomi Y. Yates, Bennett McDonald

**Affiliations:** Rural Health Services, Inc., Aiken, SC USA; Kaiser Permanente Georgia, Atlanta, GA USA

**Keywords:** antihypertensive agents, pharmacists, ambulatory care, African Americans, hypertension, multidisciplinary

## Abstract

**Introduction:** Current guidelines recommend thiazide/thiazide-type diuretics and dihydropyridine calcium channel blockers as first-line agents in treating African Americans with hypertension. The purpose of this study is to examine the impact that ambulatory care clinical pharmacy specialists (CPSs) had on initiation of first-line antihypertensives in the African American population within an integrated healthcare system.

**Methods:** This retrospective, matched, observational cohort analysis included African American patients with hypertension not receiving a first-line antihypertensive as of September 1, 2021. Patients followed by CPSs were matched up to 1:4 on age and sex to patients not followed by CPSs. The primary outcome was the percentage of patients started on first-line antihypertensive(s) after working with CPSs. Conditional logistic regression was used to analyze outcomes.

**Results:** A total of 865 patients followed by CPSs were matched to 3,192 patients not followed by CPSs. Patients followed by CPSs were initiated on first-line antihypertensives at a significantly higher rate (adjusted OR 1.98, 95% CI 1.63-2.41), and a clinically significant improvement in blood pressure was observed with systolic improving an average of 22 points and diastolic 13 points. 283 patients managed by CPSs achieved blood pressure less than 135/85 mmHg.

**Conclusion:** The initiation of first-line antihypertensives in African American patients by CPSs led to clinically significant reductions in systolic and diastolic blood pressure which supports CPS involvement in hypertension management. The assessment of study outcomes provides guidance to clinical decision-making and contributes to the development of key practice standards across both physician and clinical pharmacy specialist services.

## Introduction

Hypertension affects around 119 million adults in the United States. Approximately 79% of this population is considered to have uncontrolled blood pressure. ^1^ Multiple factors including physical inactivity, poor diet, medication nonadherence, socioeconomic status, cardiovascular comorbidities, and even racial disparities can contribute to inadequate blood pressure control.^2,3,4,5^ Uncontrolled or untreated hypertension can significantly increase the risk of end-organ damage in affected individuals.^3^ Previous studies have shown that hypertension is more prevalent in the African American population, putting them at risk for complications associated with cardiovascular disease such as heart attack and stroke.^2,5,6,7,8^

The 2017 American College of Cardiology (ACC)/American Heart Association (AHA) Clinical Practice Guidelines and 2020 International Society of Hypertension Global Practice Guidelines recommend angiotensin converting enzyme (ACE) inhibitors, angiotensin receptor blockers (ARB), thiazide/thiazide-type diuretics, and dihydropyridine calcium channel blockers (DHP-CCB) as first-line agents in the treatment of hypertension.^4^ However, guideline recommendations differ slightly for patients of African American race. African Americans have been found to have less active renin-angiotensin systems and therefore decreased blood pressure lowering with the use of ACE inhibitors and ARBs.^3,7,8^ Based on this observation, first-line treatment recommendations for African American patients include thiazide/thiazide-type diuretics and calcium channel blockers due to their effectiveness in lowering blood pressure.^4,7^

Studies have shown that collaboration between physicians and ambulatory care pharmacists results in greater blood pressure control and decreased disparities based on race and socioeconomics.^9^ A prior study conducted by Anderegg and colleagues evaluated whether pharmacist intervention could improve blood pressure in high-risk racial and socioeconomic subjects resulting in reduced healthcare disparities.^10^ In their study, pharmacists conducted comprehensive reviews with patients via telephonic and face-to-face visits. Anderegg and colleagues observed a reduction in racial and socioeconomic disparities through pharmacist-drafted care plans and physician-adjusted therapy.^10^ However, while previous studies have shown pharmacist interventions result in decreased disparities based on race and socioeconomics, limited data exists on pharmacist impact on the use of first-line antihypertensives in the management of hypertension in the African American population.^11^ The purpose of this study was to examine the initiation of first-line antihypertensive medications by ambulatory care clinical pharmacy specialists (CPSs) in the African American population within an integrated healthcare system. We hypothesized that CPSs would initiate first-line antihypertensive therapies in African American patients with hypertension at a greater rate than usual care and in turn improve overall blood pressure control in this patient population.

## Methods

### Study Setting

Kaiser Permanente Georgia (KPGA) currently serves over 320,000 members throughout twenty-six outpatient medical office buildings in the metro Atlanta area. Within the medical office buildings, CPSs are utilized in a variety of settings for ambulatory care services including but not limited to anticoagulation, cardiac risk/diabetes, chronic kidney disease, geriatrics/palliative care, heart failure, and hypertension. This study was a retrospective, observational cohort analysis conducted within the KPGA region that focused on the newly developed hypertension service that launched September 2021.

The pharmacy hypertension service is a centralized service provided by ambulatory care CPSs to optimize care for patients with hypertension by supplementing care provided by the primary care physician (PCP). Patients with a hypertension diagnosis and last office visit blood pressure reading greater than 140/90 mmHg are referred by PCPs for CPS management, as target blood pressure in clinic is considered less than 140/90 mmHg at KPGA. Pharmacy assistants outreach to schedule CPS telephone appointments for referred patients as well as for patients meeting service criteria via routine reporting. Patients were originally excluded from pharmacy management if glomerular filtration rate (GFR) was less than 30 mL/min/1.73 m^2^, if they were pregnant or receiving hospice or palliative care, or if they were prescribed hydralazine, clonidine, or labetalol medications. However, after four months of the newly launched hypertension service, criteria for the service was expanded to include patients who were prescribed hydralazine, clonidine, and labetalol medications.

Pharmacists in this service work remotely to manage blood pressure under a collaborative practice agreement. Patients enrolled in the remote hypertension service are provided blood pressure monitors at no cost then followed by ambulatory care CPSs for hypertension management. The goal of the pharmacy hypertension service is to get patients’ blood pressure controlled then transition the patients back to usual care. Goal blood pressure was considered less than 135/85 mmHg, as home blood pressure of 135/85 mmHg is equivalent to a clinic blood pressure of less than 140/90 mmHg according to the ACC/AHA guideline.^4,12,13^

Patients’ remote blood pressure results are shared electronically, with CPS appointments every two to six weeks. At service launch, patients were followed for a maximum of three to four visits before care was referred back to the PCP. However, three months after service launch, the workflow was expanded to allow up to 6 patient-pharmacist interactions before referring care back to the PCP.

### Study Design

An extensive administrative database was used to identify KPGA members diagnosed with hypertension before August 30, 2022. Patient administrative data was assessed approximately one year after the launch of the hypertension service to determine eligibility for study inclusion. African American patients with hypertension on no first-line antihypertensive followed by ambulatory care CPSs were matched to African American patients with hypertension on no first-line antihypertensive not followed by the ambulatory care CPSs. Patients were then followed for the duration of the study (September 1, 2021 to September 30, 2022), graduation from the remote pharmacy hypertension service, or until termination of their KPGA membership, whichever came first. All aspects of this study were reviewed and approved by the health system’s Institutional Review Board (IRB) before data was collected.

### Study Population

Eligible patients were self-reported African American adults (age 18 years or older) diagnosed with hypertension who had not been prescribed a first-line antihypertensive (thiazide/thiazide-type diuretic or calcium channel blocker) between September 1, 2021 and September 30, 2022. Patients were excluded if their last office visit blood pressure was less than 140/90 mmHg, GFR was less than 30 mL/min/1.73 m^2^, if they were pregnant, receiving hospice or palliative care, or had a documented intolerance to thiazide/thiazide-type diuretics or calcium channel blockers.

### Study Outcomes

The primary outcome of this study was to compare the percentage of African American patients with hypertension started on first-line antihypertensive medications who were followed by ambulatory care CPSs to those who were not followed by ambulatory care CPSs. The study also had two secondary outcomes. The first secondary outcome was to assess the percentage of African American patients that achieved home blood pressure less than 135/85 mmHg after working with ambulatory care CPSs. The other secondary outcome was to determine the change in blood pressure of African American patients after working with ambulatory care CPSs.

### Data Analysis

All patients meeting inclusion and exclusion criteria were included in the study. Covariate data was extracted from patients’ electronic medical record (EMR). Demographic characteristics used to match cases to controls included age at study start and sex. Clinical characteristics measured included vital signs such as blood pressure, BMI, and renal function. Further, comorbidities diagnosed prior to study start such as obstructive sleep apnea (OSA), diabetes, heart failure, atherosclerotic cardiovascular disease (ASCVD), and pulmonary disease were assessed via ICD codes.

African American patients with hypertension on no first-line antihypertensive followed by ambulatory care CPSs were matched up to 1:4 based on age and sex to African American patients with hypertension receiving usual care. Means were reported for interval- and ratio-level variables (e.g., blood pressure, weight, etc.) and proportions for categorical data (e.g., sex, comorbidities, etc.). McNemar’s tests were used to assess for differences in the outcome for bivariate analysis of categorical variables, and paired t-tests were used to assess for differences in the outcome for continuous variables. For the primary outcome, conditional logistic regression was utilized to compare the association between CPS interaction and usage of first-line antihypertensive medications. Unadjusted and adjusted models controlling for clinical and behavioral characteristics were reported. Secondary outcomes were assessed using descriptive analyses. Results of this study employed an intent-to-treat analysis. If patients were lost to follow up by the hypertension service, they were still included in the data analysis. All statistical analyses were performed using SAS version 8.2 Update 5.

## Results

A total of 4,943 African American patients were identified as having a documented diagnosis of hypertension prior to August 30, 2022. After screening patients for eligibility, 4,057 patients were included in the final sample size (Figure 1). Baseline characteristics are outlined in Table 1. Both groups were primarily female patients (~68%) with a mean age of 52 ± 11 years. Patients followed by ambulatory care CPSs had a greater percentage of comorbid condition diagnoses such as OSA, diabetes, ASCVD, and pulmonary disease. As seen in Table 2, when evaluating the primary outcome, patients followed by ambulatory care CPSs were prescribed first-line antihypertensives at a statistically significantly higher rate compared to those who were receiving usual care (82.4% vs. 70.8%) [Unadjusted OR 1.95, 95% CI 1.61-2.37]. This relationship remained significant in adjusted models controlling for clinical and behavioral characteristics [Adjusted OR 1.98, 95% CI 1.63-2.41].

**Table 1.** Baseline Patient Characteristics by Hypertension Management Provider.

| Characteristic | Patients Followed by<br>CPSs<br>(N = 865) | Patients Not Followed by<br>CPSs<br>(N = 3192) | p-value |
| --- | --- | --- | --- |
| Mean Age (years ± SD) | 52.5 ± 11.3 | 51.7 ± 11.2 | 0.058 |
| Female (n, %) | 598 (69.1) | 2124 (66.5) | 0.150 |
| Mean BMI (m²) | 35.1 (8.0) | 34.3 (8.5) | 0.043 |
| Comorbidity Diagnosis<br>(n, %) |  |  |  |
| Sleep apnea | 133 (15.4) | 319 (10.0) | < 0.001 |
| Diabetes | 245 (28.3) | 781 (24.5) | 0.021 |
| Heart failure | 37 (4.3) | 177 (5.6) | 0.139 |
| ASCVD | 45 (5.2) | 159 (5.0) | 0.792 |
| Pulmonary disease | 71 (8.2) | 198 (6.2) | 0.036 |
| Concomitant NSAID | 346 (40) | 1141 (35.8) | 0.021 |
| Pain Management Referral | 17 (2.0) | 55 (1.7) | 0.632 |
| Thiazide/Thiazide-type<br>Allergy | 39 (4.5) | 70 (2.2) | 0.002 |
| CCB Allergy | 74 (8.6) | 106 (3.3) | < 0.001 |

Abbreviations: CPS – Clinical Pharmacy Specialist; SD – Standard Deviation; BMI – Body Mass Index; ASCVD – Atherosclerotic Cardiovascular Disease; NSAID – Nonsteroidal Anti-inflammatory Drug; CCB – Calcium Channel Blocker

**Table 2.** Percentage of African American Patients Started on First-Line Antihypertensives.

|  | Patients Followed by<br>CPSs<br>(N = 865) | Patients Not Followed by<br>CPSs<br>(N = 3192) | Adjusted OR<br>(95% CI) |
| --- | --- | --- | --- |
| First-Line Antihypertensive<br>Initiated<br>n (%) | 713 (82.4) | 2259 (70.8) | 1.98 (1.63-2.41) |

Abbreviations: CPS – Clinical Pharmacy Specialist; OR – Odds Ratio; CI – Confidence Interval

**Figure 1.**
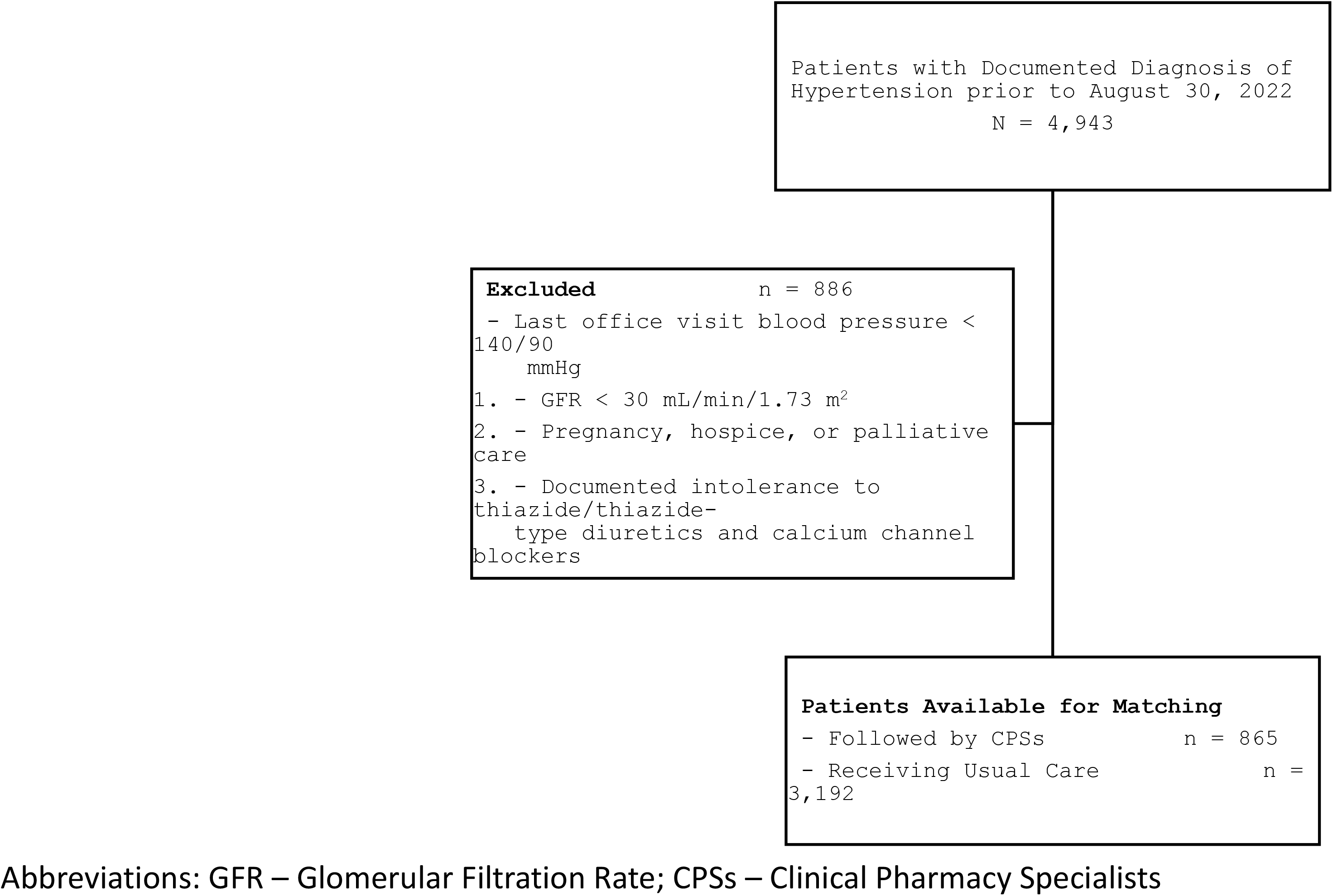
African American Participants Eligible for Study Inclusion.

Thirty-three percent of patients working with ambulatory care CPSs achieved blood pressure less than 135/85 mmHg (Figure 2). Additionally, systolic blood pressure improved on average 22 points and diastolic blood pressure improved 13 points after working with ambulatory care CPSs (Figure 3).

**Figure 2.**
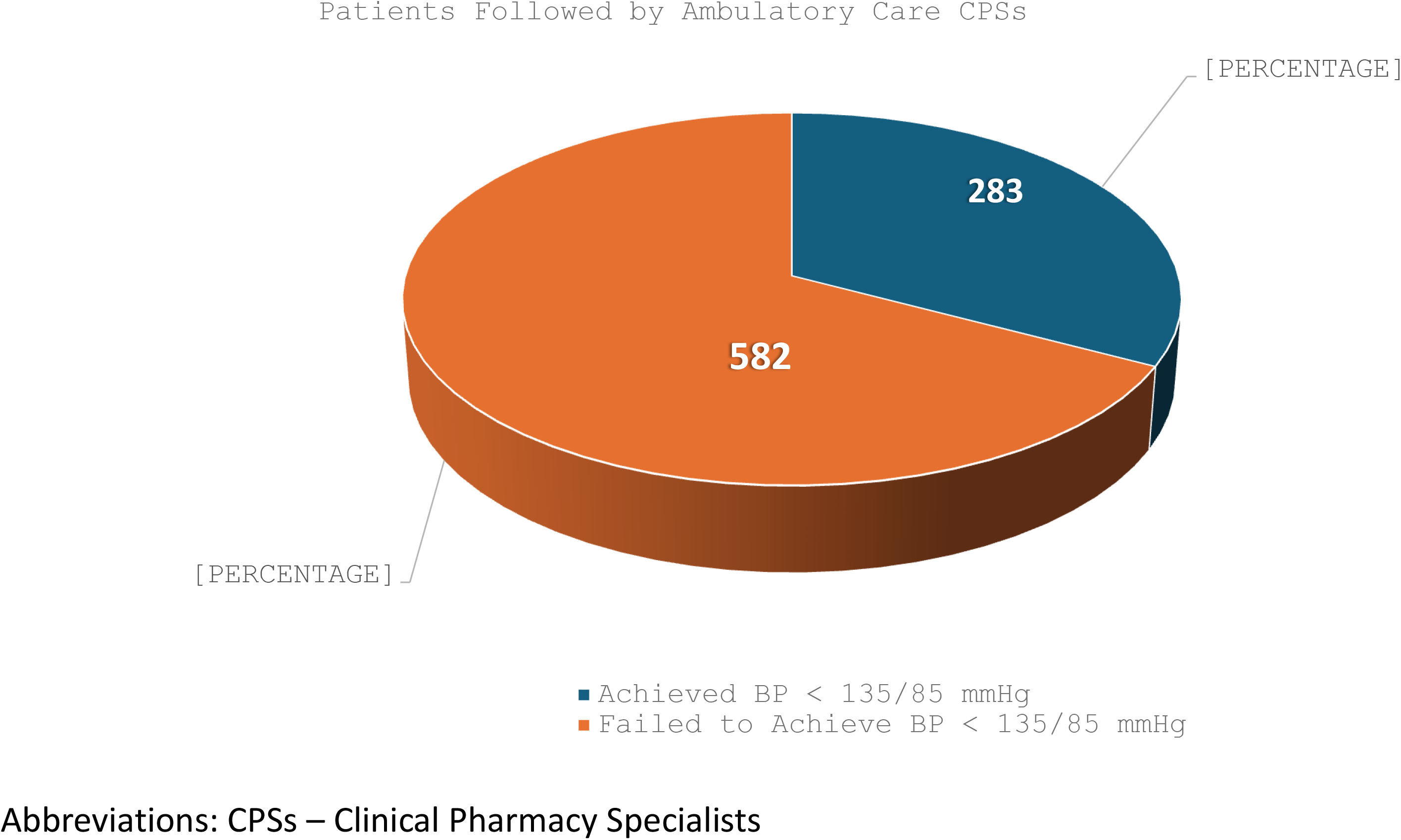
Percentage of Patients who Achieved versus Failed to Achieve Blood Pressure Goal.

**Figure 3.**
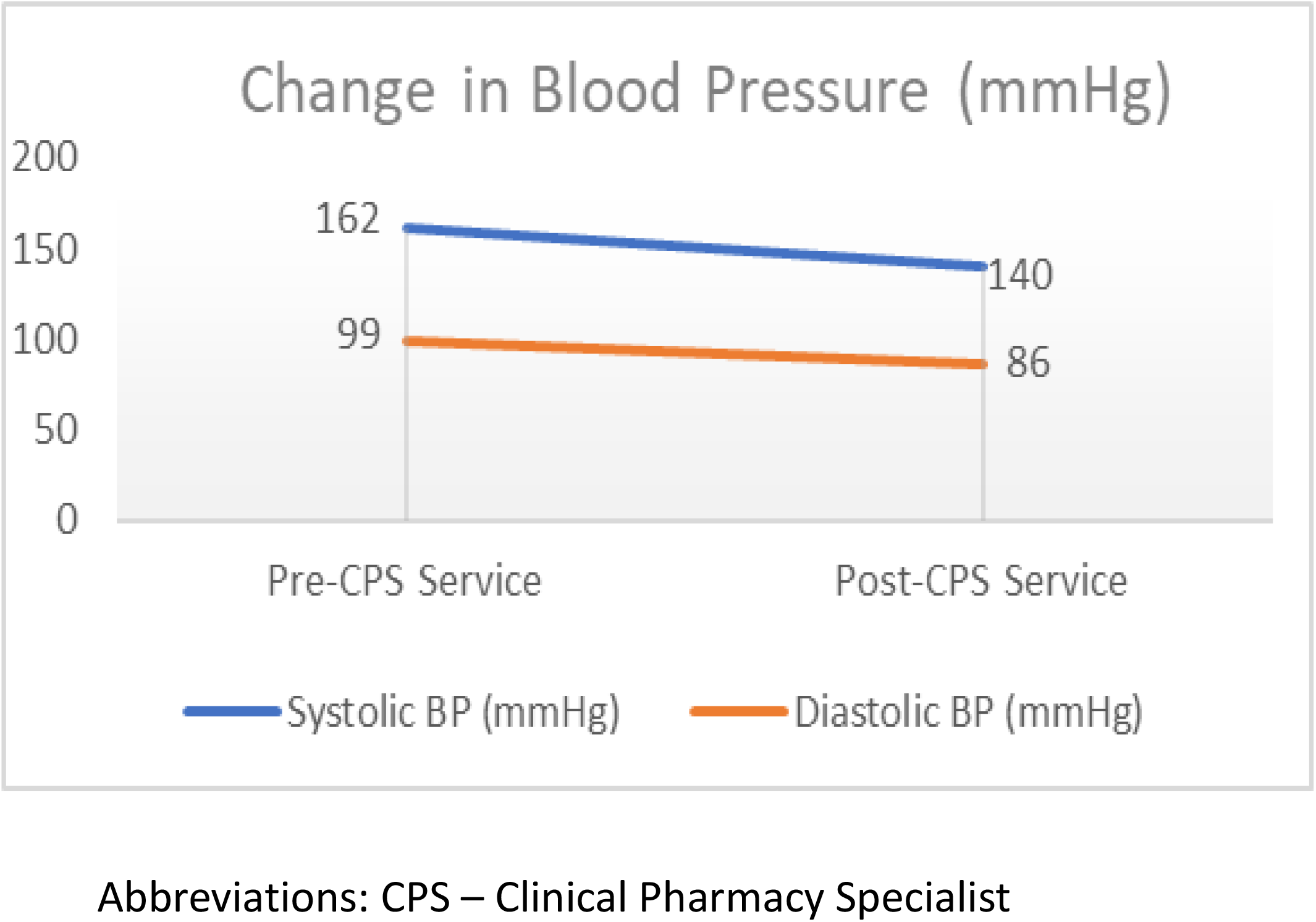
Change in Blood Pressure After Working with Ambulatory Care CPS.

## Discussion

In this retrospective, matched, observational cohort analysis, a higher percentage of African American patients were prescribed guideline recommended first-line antihypertensives when followed by ambulatory care CPSs in comparison with usual care. A previous study by Ettehad and colleagues suggests that for every 10 mmHg reduction in systolic blood pressure, a patients’ risk of major cardiovascular disease events, coronary heart disease, and heart failure is significantly reduced.^14^ Despite the majority of patients not achieving goal blood pressure in our study, a clinically significant improvement was observed when assessing change in blood pressure in patients followed by ambulatory care CPSs. The lower-than-expected percentage of patients achieving goal blood pressure could be attributed to the newness of the service, limited pharmacist interactions directly after the service launch, and/or a greater percentage of patient comorbidities such as OSA and diabetes in the CPS study arm.

Our findings on blood pressure improvement are similar to those of the trial conducted by Anderegg and colleagues. The study’s main results showed that the difference in mean systolic and diastolic blood pressure between the intervention and usual care groups was −6.1 and −2.9 mm Hg at 9 months (p<0.001 and p=0.003, respectively).^10^ Compared to this previous study, our study observed greater mean systolic blood pressure lowering of 22 points and diastolic blood pressure lowering of 13 points on average after up to six patient-pharmacist interactions. In addition to observing greater reductions in blood pressure, our study differed by leveraging remote technology to monitor patient’s home blood pressure. This is novel and important to evaluate considering that African American patients are more likely to experience masked hypertension, where readings that appear acceptable in office run higher at home.^7^ Given the availability of the remote data, pharmacists had the opportunity to provide appropriate counseling and adjust medications accordingly during each interaction. In addition to utilizing remote technology, our study included a large patient sample size through an integrated healthcare system and was able to examine the initiation and usage of first-line antihypertensive agents in African American patients.

While our study had several notable strengths, there were some limitations. The first limitation was that our data emanated from a new clinical pharmacy service. After a five-month pilot, the hypertension service fully launched in September 2021 which was also the start of our study period. Additionally, the quantity of CPS interactions differed throughout the study period as more CPS resources were obtained. As the service grew, CPS interactions per patient increased from up to three to up to six interactions before patients were referred back to usual care. Additional research could be performed for further assessment of results including CPS impact on medication adherence or the impact on occurrences of hypertensive urgency or emergency within our patient population.

The initiation of first-line antihypertensives in African American patients by ambulatory care CPSs within this integrated healthcare system led to clinically significant decreases in systolic and diastolic blood pressure. These reductions in blood pressure support CPS involvement in hypertension management. The assessment of study outcomes provides guidance to clinical decision-making and contributes to the development of key practice standards across both physician and clinical pharmacy specialist services.

## Data Availability

An extensive, internal, administrative database not accessible to the public was used to identify Kaiser Permanente (KP)- Georgia members diagnosed with hypertension before August 30, 2022. All data will remain internal on KP servers that is password protected and patients will be only identifiable by medical record number. PHI will be destroyed at the earliest opportunity after publication of the findings is complete; no data will be stored for future research.

## Acknowledgements

This work would not be possible without the collective efforts from our colleagues at Kaiser Permanente who provided insight and expertise that greatly assisted the research. We thank Zuwere Bailey, Business Intelligence Analytics Consultant for her assistance with data collection and Bennett McDonald, Programmer/Analyst II for his assistance with statistical analysis. We would also like to thank Thomas Delate for sharing his expertise throughout the study design and manuscript revision process. Their assistance greatly strengthened this research project.

